# COVID-19 healthcare demand and mortality in Sweden in response to non-pharmaceutical (NPIs) mitigation and suppression scenarios

**DOI:** 10.1101/2020.03.20.20039594

**Authors:** Henrik Sjödin, Anders F. Johansson, Åke Brännström, Zia Farooq, Hedi Katre Kriit, Annelies Wilder-Smith, Christofer Åström, Johan Thunberg, Mårten Söderquist, Joacim Rocklöv

## Abstract

**Background:** While the COVID-19 outbreak in China now appears surpressed, Europe and the US have become the epicenters, both reporting many more deaths than China. Responding to the pandemic, Sweden has taken a different approach aiming to mitigate, not suppressing community transmission, by using physical distancing without lock-downs. Here we contrast consequences of different responses to COVID-19 within Sweden, the resulting demand for care, intensive care, the death tolls, and the associated direct healthcare related costs.

**Methods:** We use an age stratified health-care demand extended SEIR compartmental model calibrated to the municipality level for all municipalities in Sweden, and a radiation model describing inter-municipality mobility.

**Results:** Our model fit well with the observed deaths in Sweden up to 20^th^ of April, 2020. The intensive care unit (ICU) demand is estimated to reach almost 10,000 patients per day by early May in an unmitigated scenario, far above the pre-pandemic ICU capacity of 526 beds. In contrast, a scenario with moderate physical distancing and shielding of elderly in combination with more effective isolation of infectious individuals would reduce numbers to below 500 per day. This would substantially flatten the curve, extend the epidemic period, but a risk resurgence is expected if measures are relaxed. The different scenarios show quite different death tolls up to the 1^th^ of September, ranging from 5,000 to 41,000 deaths, exluding deaths potentially caused by ICU shortage. Further, analyses of the total all-cause mortality in Stockholm indicate that a confirmed COVID-19 death is associated with a additional 0.40 (95% CI: 0.24, 0.57) all-cause death.

**Conclusion:** The results of this study highlight the impact of different combinations of non-pharmaceutical interventions, especially moderate physical distancing and shielding of elderly in combination with more effective isolation of infectious individuals, on reducing deaths and lower healthcare costs. In less effective mitigation scenarios, the demand on ICU beds would rapidly exceed capacity, showing the tight interconnection between the healthcare demand and physical distancing in the society. These findings have relevance for Swedish policy and response to the COVID-19 pandemic and illustrate the importance of maintaining the level of physical distancing for a longer period to suppress or mitigate the impacts from the pandemic.

**Key messages:**

- We find physical distancing and isolation of infectious individuals without lockdown is effective in mitigating much of the negative direct health impact from the COVID-19 pandemic in Sweden, but has a higher death toll compared to other Scandinavian countries who did implement a lockdown
- Between the start of the Swedish model of physical distancing and shiedling the elderly in March to late April, it appears Sweden has managed to ensure that ICU demands do not exceed ICU capacities and that deaths are substantially reduced compared to a counterfactual scenario.
- In the counterfactual scenario (eg no public health interventions), the intensive care unit demand is estimated to be almost 20 times higher than the intensive care capacity in Sweden and the number of deaths would be between 40,000 to 60,000
- Under current mitigation strategies, the death toll, health care need, and its associated cost are, however, still substantial, and it is likely to continue to rise unless the virus is suppressed, or eliminated. In the stronger mitigation and suppression scenarios, including the scenario fitting best to data from Sweden by late April 2020, there is an obvious risk of resurgence of the epidemic unless physical distancing, shielding of the elderly, and home isolation are effectively sustained.
- Additional analyses indicate all-cause non COVID-19 excess mortality rises with 0.4 deaths per every reported COVID-19 death in the Stockholm area.

## Introduction

The novel SARS-CoV-2 is highly transmissible(1), rapidly spread around the globe since it emerged in Wuhan, China(2), at a rate much faster than other emerging infectious diseases such as Ebola.(3) In response to the COVID-19 outbreak, China implemented extraordinary public health measures at great socio-economic cost, moving swiftly to ensure early identification of cases, prompt laboratory testing, facility-based isolation of all cases, contact tracing and quarantine.(4) In the community, physical distancing was implemented at a grand scale, all mobility put to an halt, and the city of Wuhan was in lock-down for about 9 weeks.(5) China’s tremendous efforts showed success.(6) Other Asian countries facing a major explosion such as South Korea also managed to curb the epidemic. South Korea employed very liberal testing, hospital-based isolation of all cases, combined with extensive contact tracing enhanced by mobile phone and digital technologies, but did not use a lock-down.(7, 8)

While the outbreak in China appears to be contained, since mid March 2020, the epicenter of the COVID-19 pandemic is in Europe, and since April in the United States. There is thus an urgent need to determine how best to reduce transmission rates, the height of the epidemic peak, the peak demand on healthcare services, and how to reduce fatalities.

In the absence of vaccines, a wide range of control measures can be considered to contain or mitigate COVID-19. These include active case finding with prompt isolation of cases, contact tracing with quarantine of contacts, school closures and closures of public places, mobility restrictions, physical distancing in the community, physical distancing only of the elderly, and a lock-down (also known as Cordon sanitaire).(4) There is currently no consensus about which measures should be considered, in which combination, and at which epidemiological threshold such measures should be implemented for maximum public health impact.(9)

Two strategies can be considered: (a) suppression which aims to rapidly reverse epidemic growth, thereby reducing case numbers to low levels, and (b) mitigation, which focuses on slowing but not necessarily immediately stopping epidemic spread – reducing peak healthcare demand while shielding those most at risk of severe disease from infection. Each policy has major challenges. Suppression aims to rapidly reduce the reproduction number, *R*_0_, to below 1, thus causing case numbers to consistently decline. Mitigation aims to slow spread by reducing *R*_0_ to a value close to but slightly above 1 for some time before the epidemic growth will cease gradually, partially in reponse to increasing levels of disease immunity in the population.

Public health measures need to be weighed up against economic repercussions and mental health of prolonged lock-downs. Although more strict community containment measures such as lock down will result in a shorter duration of the outbreak(10), the non-health sector negative consequences may be huge.

Sweden decided to implement public health interventions without a lockdown. Schools and universities were not closed, restaurants and bars remained open, instead Swedish citizens implemented "work from home" policies where possible, social distancing without police enforcement, and shielding of those older than 65 of age.

Here we aim to quantify the effects of the Swedish measures. We estimate the impact of COVID-19 on the Swedish population at the municipality level, considering demography and human mobility under various scenarios of mitigation and suppression. We estimate the time course of infections, health care needs, and the mortality in relation to the Swedish ICU capacity, as well as the costs of care, and compared alternative policies and counterfactual scenarios.

## Methods

We developed a compartmental epidemiological model based on the SEIR formulation, and extended it to account for additional variables including compartments for health and ICU care. All these variables were age-structured (0-59, 60-79, and 80+ years). The model included age-structured compartments for susceptibles, exposed, infected, inpatient care, ICU care, dead and recovered populations based on Swedish population data at the municipality level (see Supplementary Information 1). Overall, the population of infected individuals were divided into two different groups, those that had sufficiently severe symptoms to potentially end up in hospital care (1 out of 6), and those who had mild or asymptomatic infections or were sick at home (5 out of 6).(11, 12) This parameter was calibrated to data. The model allowed for three different ways that deaths could occur: 1) after unsuccesful ICU treatment; 2) after triage and denial of ICU due to low chances of surviving or when ICU demand exceeds ICU capacity; and 3) outside of healthcare.

The model captured spatial demographic heterogeneities at the level of municipality in Sweden, and inter-municipality travelling based on a radiation model (see Supplementary Information 1). The radiation model was calibrated using a N1H1 Influenza A model and data for the period 2015-2018 in Sweden. Demographic data was obtained at the municipality level for the year 2018 from Statistics Sweden.

The parameterization of the model was achived in to ways: (1) some parameters were set to fixed values based on what is known through literature and data; (2) some parameters were given initial values from the international literature and then calibrated by fitting the model to current available outbreak-data on infection prevalence, deaths, ICU load and healthcare in Sweden. The full set of parameter values are given in the Supplementary Information Table S1.1. Age specific health-care need parameters from Ferguson et al. was initially used to represent the three age groups in our study (Supplementary Information 1, Table S1.2). These values where then calibrated to better reflect observed infection prevalence, in-patient care demand, ICU care demand and registered deaths due to COVID-19 from Stockholm region and Sweden (See Supplement S1.2).

The infectious period is likely to vary by the individual and range from days to weeks. Viral shedding is reported to occur from 7 to 22 days, including in mild cases of disease (13), and is a driver of disease transmission. Isolation of patients, or staying home if presenting with symptoms, will reduce transmission to contacts, and is a key strategy to contain COVID-19. It shortens the period that infected persons are able to infect others. Importantly, transmission from an infected but asymptomatic or pre-symptomatic individuals can still occur despite control measures.(14) We assumed that the average effective infectious period in the general population is 5 days, shortened from 7 days by natural isolation of symptomatic infected individuals. We assumed that individuals going into healthcare were admitted on average after 3 days of symptomatic infection, and were isolated from transmitting the virus to other individuals while in hospital care.

We used a measurement from the Swedish Public Health Agency of virus prevelence measured by nucleic acid detection in nasopharyngeal samples from the general population in the Stockholm region centered around the 1th of April. The measurement indicated 2.5% (95 % Cl = 1,4 – 4,2 %) of the population were infected.(15) To calibrate the infection prevalence in the model with the study result, we adjusted the model by allowing a latent compartment after infectiousness for the general infected population. Our model thus takes into account that virus ion can be detected for 7 days.

We calibrated the daily transmission rate, *β*, to 0.91*c*(*t*) to Swedish data on healthcare load, virus prevalence, and mortality (Supplementary Information 1), where *c*(*t*) is a time dependent contact-rate scaling parameter (Supplementary Information 1). In our model, *R*_0_ is dependent on contact-rate and infectious period which parameters in turn are dependent on age-distribution (See Supplementary Information 1). Accordingly, we account for spatial and demographical heterogeneity in *R_0_.* The country-level *R*_0_ is the average over all municipality-local values for *R_0_.* It is not with in the scope of the paper to derive exact values for *R_0_,* yet we can see that the within-municipality *R_0_,* for the no-countermeasures scenario (i.e., scenario a), is within the range of 2.73 to 4.55, and this range is consistent with the reported basic reproduction rates for COVID-19.(16) The country-level *R*_0_ is given by an average over municipality-local *R*_0_ values. Due to the many travelers infected with SARS-CoV-2 arriving from Italy, Austria and other parts of Europe, in the week of the 24^th^ of February, the model was seeded with 1 case per 120,000 individuals for all municipalities except for the municipalities within Region Stockholm that were seeded with 1 case per 30,000 individuals.

The model was set up to predicting the municipality transmission dynamics and inter-municipality spread across Sweden starting from 24^th^ of February and ending a little more than 6 months later, September 1, 2020. The model used scenarios to describe the countermeasures and counterfactual impacts. The mitigation and suppression scenario had onset the 20^th^ of March by a transient function with full effect by the end March (Supplementary Information 1, Table S1.1). The five different scenarios is summarized by:

a. no public health interventions (counterfactual scenario);
b. modest physical distancing in ages 0-59 years, moderate in ages 60+ years;
c. modest physical distancing in ages 0-59 years, moderately strong in ages 60+ years;
d. moderate physical distancing in ages 0-59 years, strong in ages 60+ years, and with an inceased degree of isolation of infectious individuals;
e. same as in scenario d), but with a slightly higher degree of isolation of infectious individuals.

A complete description of the scenarios is provided in the Supplementary Information 1.

In the scenarios the ICU capacity were compared against current baseline availability of 526 ICU beds in Sweden, which has been doubled in respons to the COVID-19 pandemic. The numbers of deaths and the infection fatality rate (IFR) associated with the different mitigation and supression scenarios were derived from our model. We also derived the direct healthcare cost for each of the scenarios (see Supplementary Information 4).

Additionally, we extracted total all-cause of deaths from Stockholm region and estimated a potential excess mortality beyond confirmed COVID-19 cases. This was done by comparing the excess mortality during the COVID-19 outbreak with the mortlity the weeks before the outbreak for the same and previous years using time series regression methods, while adjusting for time trends and the patterns of deaths previous months and years (Supplementary Information 3).

## Results

The model showed a good fit against the reported COVID-19 related deaths in Sweden up to 20^th^ of April, 2020, in scenario d (Figure 1m-p). In Table 1 we present the R^2^ and the mean square error of the observations to the predictions for scenario (a) to (e) in relation to: deaths for Sweden as a whole, deaths in Stockholm, ICU bed demand in Stockholm, and inpatient care in Stockholm. Scenario (d) further well describes the observation that 2.5% of the population in Stockholm were infected with the SARS-CoV-2 virus centered around the 1 of April (Figure 2d). Overall, the IFR for Sweden in scenario (a) to (e) is estimated to 0.40; 0.40; 0.37; 0.28 and 0.27, respectively (Table 1).

**Figure 1.**
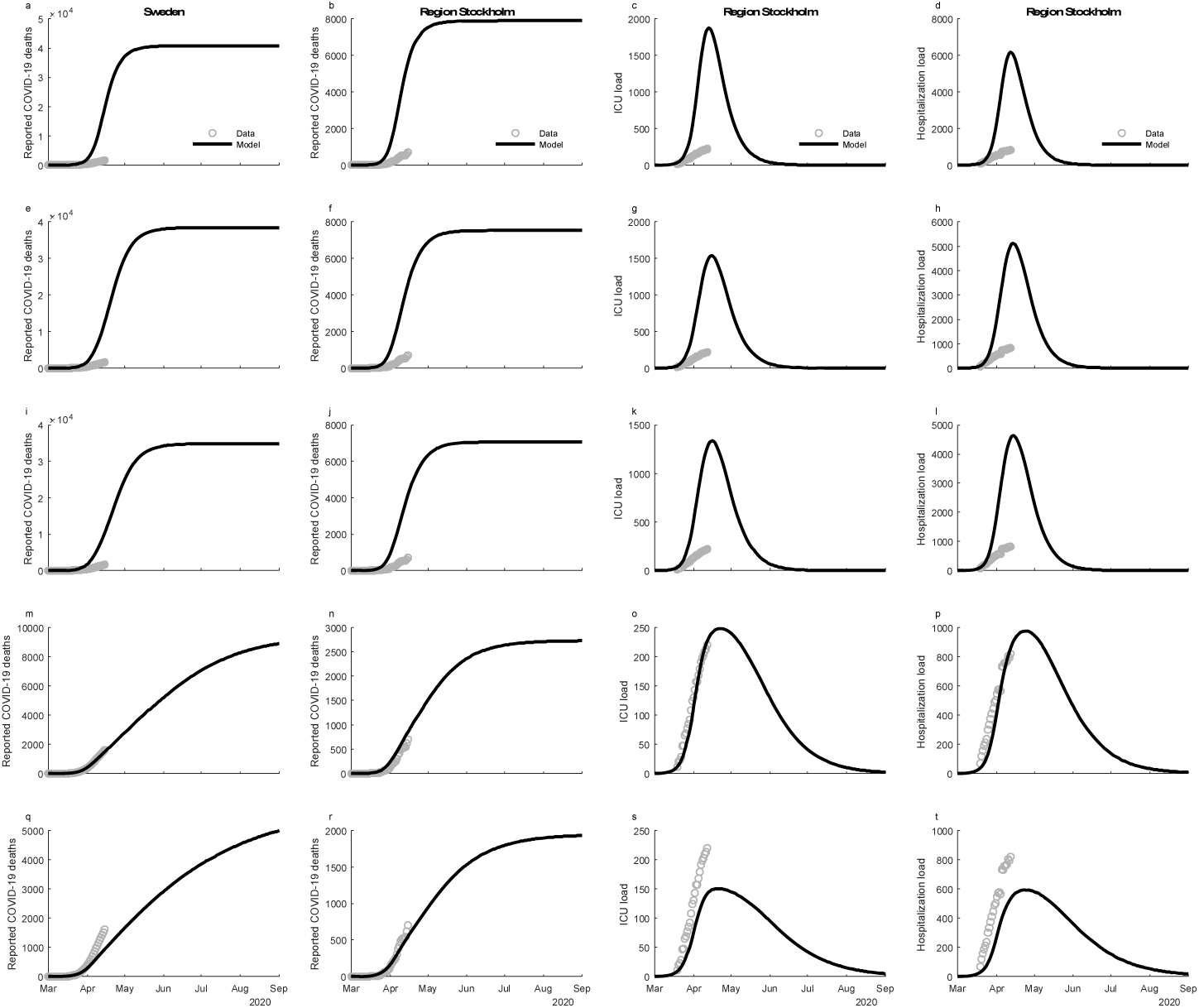
Predicted number of total deaths from COVID-19 in the whole population in Sweden (first column), and for Stockholm region (second column); predicted demand of ICU beds in Stockholm (third column), and; inpatient care beds in Stockholm (fourth column). Actual observations in the early phase of the outbreak are illustrated as circles (O). The mitigation scenarios are organized in rows with panel a) no public health interventions (counterfactual scenario); b) modest physical distancing in ages 0–59 years, moderate in ages 60+ years; c) modest physical distancing in ages 0–59 years, moderately strong in ages 60+ years; d) moderate physical distancing in ages 0–59 years, strong in ages 60+ years, and improved isolation of infectious individuals; e) moderate physical distancing in ages 0–59 years, strong in ages 60+ years, and further improved isolation of infectious individuals. Mitigation giving rise to these predicted values had onset the 20^th^ of March.

**Table 1.**
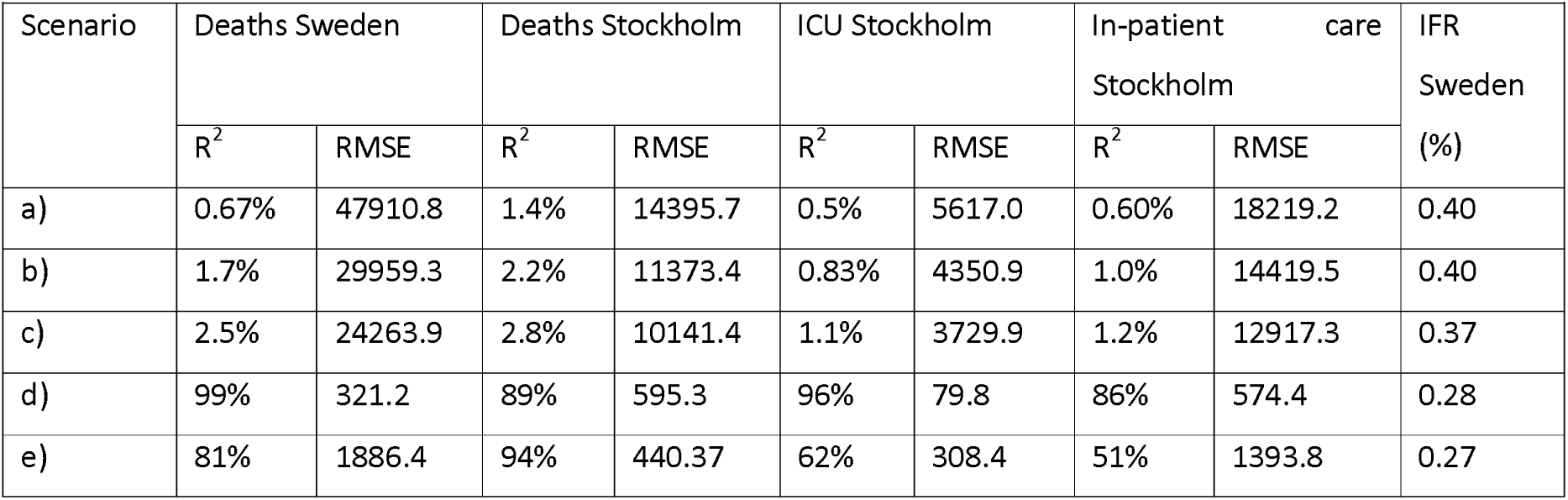
Estimates of R^2^ and root-mean-square error (RMSE) for the different model scenarios to observations from Sweden and Stockholm, along with the resulting infection fatality ratio (IFR) for Sweden.

In Figure 3 we present the scenarios of country level COVID-19 ICU bed demand over time in Sweden per the different age groups, and in total. According to scenario (a), the outbreak would peak in the middle of April and reach an ICU bed demand of around 10,000 patients (Figure 3; panel a). The age group below 60 years of age alone would take up more than the baseline ICU resources of 526 beds during a month at the peak. According to scenario (b), the ICU demand would peak at around 7,000 beds at the peak around the first of May (Figure 3; panel b). The demand would be flattened and continue for a longer period. The ICU demand for those below 60 years of age would again exceed the baseline ICU beds for 1 month. According to scenario (c), the ICU bed demand would peak at around 6,000 at the peak around the first of May (Figure 3; panel c). The demand would be flattened and continue for a longer period. The ICU bed demand for those aged below 60 years would almost take up the baseline ICU beds for a period slightly less than 1 month. According to scenario (d), the intensive care demand would not increase beyond 500, it would peak in June and decrease in August (Figure 3; panel d). According to scenario (e), the intensive care demand would be very small for the whole study period (Figure 3; panel e). We note, that the outbreak would likely resurge unless others means are there to control the transmission when the countermeasures are lifted in scenario (d) and (e). Corresponding estimates for Stockholm region are given in Figure 4 (panel a-e). Notewhorthy, the healthcare demand peaks earlier in Stockholm as compared with Sweden as a whole and decreases earlier in the summer.

**Figure 3.**
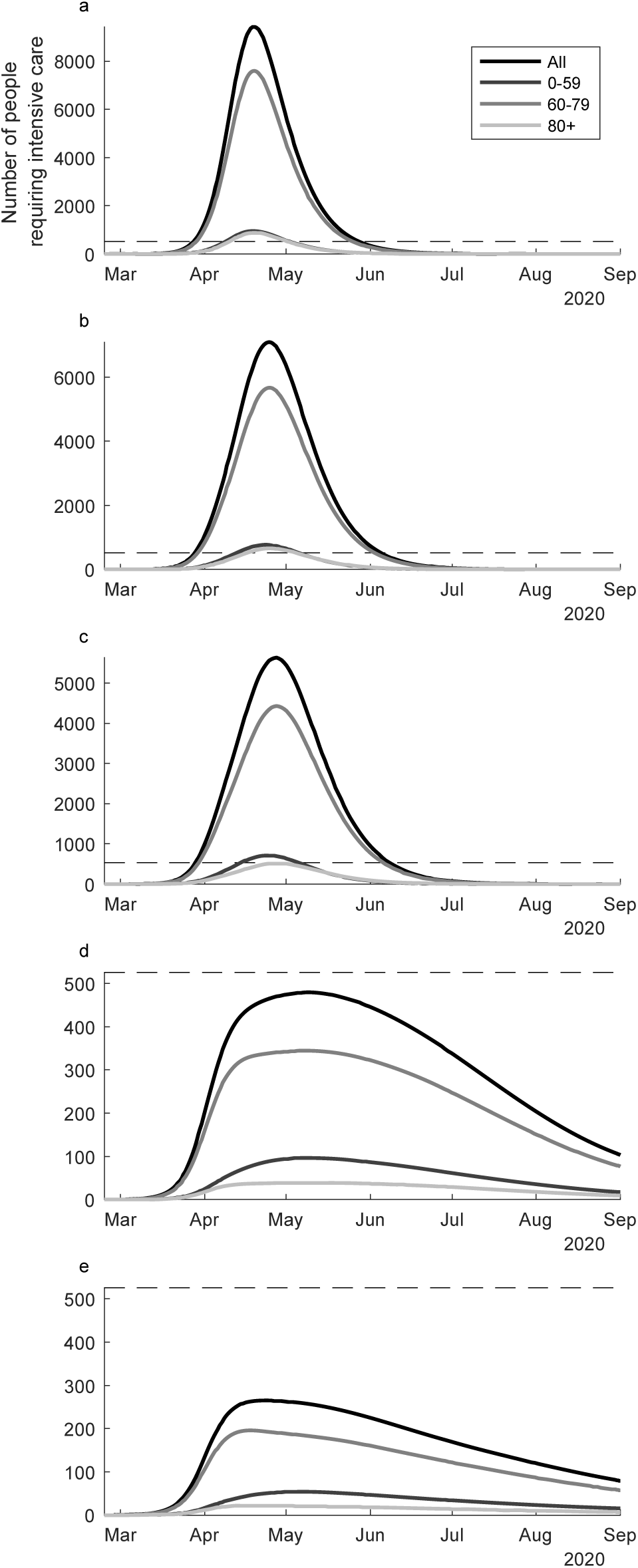
The predicted ICU bed demand per day from 24^th^ of February to the 1st of September, 2020, overall in Sweden in relation to different suppression & mitigation scenarios. Panel a) no public health interventions (counterfactual scenario); b) modest physical distancing in ages 0–59 years, moderate in ages 60+ years; c) modest physical distancing in ages 0–59 years, moderately strong in ages 60+ years; d) moderate physical distancing in ages 0–59 years, strong in ages 60+ years, and improved isolation of infectious individuals; and e) moderate physical distancing in ages 0–59 years, strong in ages 60+ years, and further improved isolation of infectious individuals. Mitigation giving rise to these predicted values had onset the 20^th^ of March.

**Figure 4.**
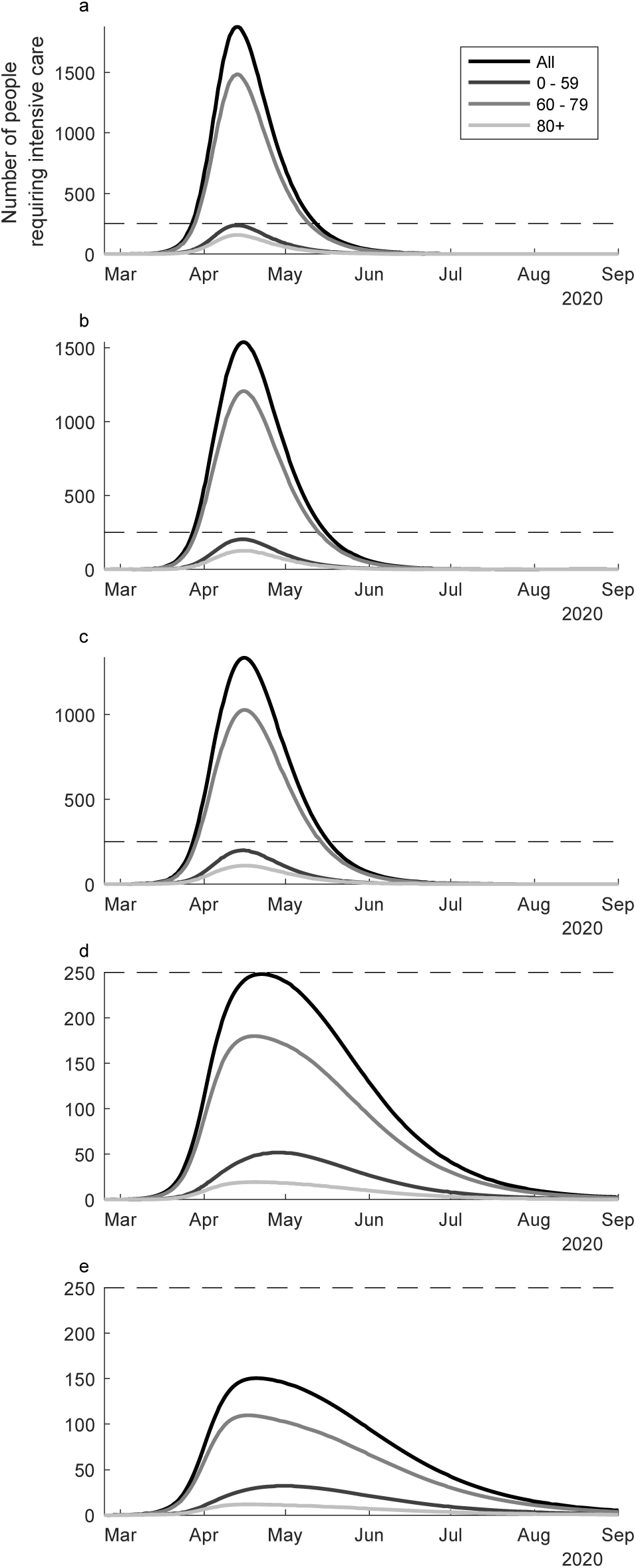
The predicted ICU bed demand per day from 24^th^ of February to the 1st of September, 2020, in the region of Stockholm in relation to different suppression & mitigation scenarios. Panel a) no public health interventions (counterfactual scenario); b) modest physical distancing in ages 0–59 years, moderate in ages 60+ years; c) modest physical distancing in ages 0–59 years, moderately strong in ages 60+ years; d) moderate physical distancing in ages 0–59 years, strong in ages 60+ years, and improved isolation of infectious individuals; and e) moderate physical distancing in ages 0–59 years, strong in ages 60+ years, and further improved isolation of infectious individuals. Mitigation changes giving rise to these predicted values had onset the 20^th^ of March.

In the Supplementary Information 2 we show how the timing of the increase in cases across the scenarios a-e presented in Figure 2 is sensitive to mobility between municipalities (Figure S2.1). There is a slightly earlier increase in ICU bed demand with higher inter-municipality mobility.

**Figure 2.**
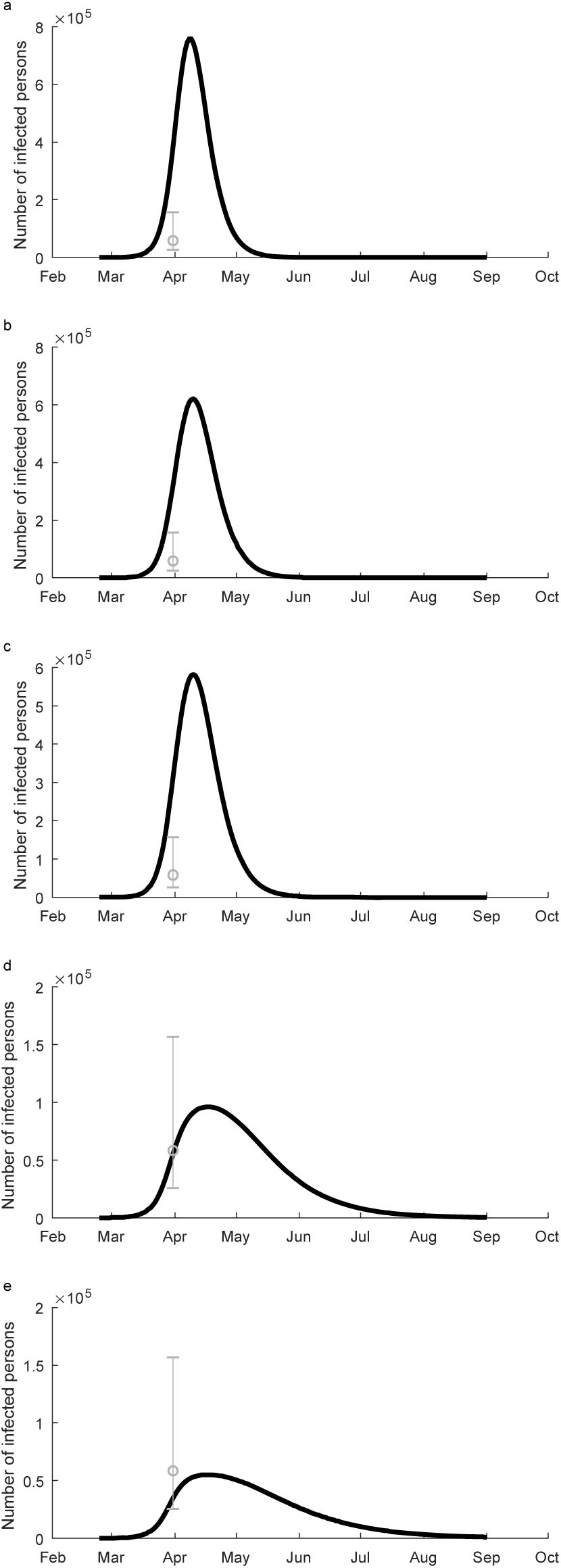
The number of individuals in in the Stockholm region population predicted to carry the virus over time as determined by a virus detection assay. An actual observed result from the 1 April, 2020, using a population sample is illustrated by a circle (O) with 95% Cl (vertical bars). Panel a) no public health interventions (counterfactual scenario); b) modest physical distancing in ages 0–59 years, moderate in ages 60+ years; c) modest physical distancing in ages 0–59 years, moderately strong in ages 60+ years; d) moderate physical distancing in ages 0–59 years, strong in ages 60+ years, and improved isolation of infectious individuals; and e) moderate physical distancing in ages 0–59 years, strong in ages 60+ years, with further improved isolation of infectious individuals. Mitigation giving rise to these predicted values had onset the 20^th^ of March.

In Table 2 we describe the predictions of the total number of individuals infected, the total person days of care, the total person days of ICU occupancy, the total number of deaths (assuming all ICU demands are satisfied), and the deaths from ICU capacity shortage (100% above baseline level). Following on this, we estimate the direct costs of the care and intensive care demands (Table 3). The number of infected individuals in Sweden is predicted very high by the model in scenario (a) to (c) with attach rates beyond 90% of the population already by the 1th of September, 2020 (Table 2). In the scenario (d) and (e), only 3.2 and 1.8 million people, respectively, would be infected by 1^th^ September.

**Table 2.** Estimates of infections and healthcare demand aggregated over Sweden for the period 24^th^ February to 1^st^ September, 2020.

| Mitigation and suppression actions | Total number of individuals infected | Person days in in-patient care | Person days in the ICU | Total number of deaths assuming all ICU demands are satisfied | Total number of deaths from ICU shortage |
| --- | --- | --- | --- | --- | --- |
| *a) no public health interventions (counterfactual scenario)* | 10,104,140 | 766,940 | 268,215 | 40,820 | 21,228 |
| *b) modest physical distancing in ages 0-59 years, moderate in ages 60+ years* | 9,694,549 | 726,716 | 251,666 | 38,436 | 18,397 |
| *c) modest physical distancing in ages 0-59 years, moderately strong in ages 60+ years* | 9,315,236 | 671,978 | 225,446 | 34,830 | 15,191 |
| *d) moderate physical distancing in ages 0-59 years, strong in ages 60+ years, and improved isolation of infectious individuals* | 3,209,317 | 189,574 | 52,804 | 8,883 | 0 |
| *e) moderate physical distancing in ages 0-59 years, strong in ages 60+ years, and further improved isolation of infectious* | 1,878,326 | 107,543 | 29,178 | 4,985 |  |
| *individuals* |  |  |  |  | 0 |

**Table 3.** Estimates direct costs of infections and healthcare demand aggregated over Sweden for the period 24^th^ February to 1^th^ September, 2020.

|  | Costs of<br>cumulative<br>person days in<br>in-patient care<br>(million<br>SEK/2020) | Costs of<br>cumulative<br>person days in<br>intensive care<br>(million<br>SEK/2020) | Total direct<br>healthcare<br>costs |
| --- | --- | --- | --- |
| Mitigation and suppression actions: |  |  |  |
| *a) no public health interventions (counterfactual scenario)* | 12,000 | 8,000 | 20,000 |
| *b) modest physical distancing in ages 0-59 years, moderate in ages 60+ years* | 11,000 | 8,000 | 19,000 |
| *c) modest physical distancing in ages 0-59 years, moderately strong in ages 60+ years* | 10,000 | 7,000 | 17,000 |
| *d) moderate physical distancing in ages 0-59 years, strong in ages 60+ years, and improved isolation of infectious individuals* | 3,000 | 2,000 | 5,000 |
| *e) moderate physical distancing in ages 0-59 years, strong in ages 60+ years, and further improved isolation of infectious individuals* | 2,000 | 2,000 | 4,000 |

Overall, the scenarios show the demand on inpatient care varies from just below 700,000 person-days to just above 100,000 person-days and the, while the demand on intensive care range from around 270,000 to around 30,000 person-days. The death rates, assuming no limits in ICU, varies from around 40,000 to 5,000 depending on the mitigation and suppression actions. Assuming instead a cap of the ICU bed capacity of 100% above baseline, the estimated number of additional excess deaths from lack of ICU capacity varies from 21,000 to 0 depending on the scenario. The total direct medical cost range between 20 billion to 4 billion SEK depending on the scenario (Table 3).

From the calculation of excess deaths associated with the COVID-19 outbreak in Stockholm (Supplementary Information 3), we observe a linear increase in all-cause deaths of 0.40 (95% CI = 0.24, 0.57) for each registered COVID-19 death. The analyses excluded reported COVID-19 deaths from all-cause mortality, and thus indicate 40% additional deaths beyond reported COVID-19 cases.

## Discussion

Our study shows an exponential growth of the number of COVID-19 infections, health care demands and deaths in Sweden which became apparent towards the end of March and the beginning of April, 2020. In April, it further suggests a strong effect of the physical distancing efforts put into place around the 20^th^ of March in Sweden. The epidemiological data from Sweden align best to our modelled secenario (d) which describe a moderate physical distancing in those below 60 years of age, a strong shielding of those above 60 years of age, and improved awareness and compliance of home isolation of symptomatic COVID-19 cases. The level of physical distancing and isolation in this scenario did not compromise the access to health care, and did not overwhelm the health care system. The extent of measures were less stringent and economically damaging compared to those introduced in other Scandinavian countries (Norway, Finland, Denmark), but the number of deaths by 5 May 2020 was much higher: according to the WHO situation report on 5 May 2020, there were 2,769 deaths with 22,721 confirmed infections in Sweden, versus 493 deaths with 9,670 reported cases in Denmark, 208 deaths with 7,847 cases in Norway, and 240 deaths with 5,327 cases in Finland.(17)

In the counterfactual scenario (eg no public health interventions), the intensive care unit demand was estimated to be almost 20 times higher than the intensive care capacity in Sweden and the number of deaths would be between 40,000 to 60,000

Our estimates using the Swedish model of physical distancing without lockdown show that by end August, 2020, about 30% of the Swedish population will be infected, resulting in about 9,000 deaths by that time point. Despite such high exposure, Sweden would remain far below the herd immunity needed to stop the outbreak, a threshold estimated to be around 60–70%). (18) Therefore, up to September 2020, the predicted impact is very dependent on ongoing adherence to physical distancing and shielding of vulnerable age groups. If such measures are not maintained, the consequences would be severe with demands substantially exceeding health care capacity and mortality rates rapidly increasing. Natural herd immunity (i.e., a situation where *R*_0_ goes below 1 even if countermeasures were discontinued) appears not a viable objective to stop the virus circulation given the predicted death tolls. Further measures, including enhanced testing, prompt isolation of cases, more effective contact tracing and quarantining of contacts(19, 20), would result in further reducing transmission intensity and daily new cases, while avoiding lockdowns, until other control options such as vaccine and effective therapeutic options are readily available.

Of note, due to the strong triage in our model with only 15% of those aged 80+ being treated at the ICU care, the demand of this group is overall already estimated to be very low. In order to capture the Swedish mortality patterns, our model further estimates a substantial number of deaths occurred outside hospitals, mainly in care homes. The excess mortality estimates published by the European Centre for Disease Control indicate a substantial amount of additional deaths must have occurred due to COVID-19. (15) The total all-cause mortality in Stockholm indicate that a confirmed COVID-19 death is associated with an additional 0.40 (95% CI: 0.24, 0.57) all-cause death eg 40% additional deaths beyond the reported COVID-19 cases.

In Sweden, we calibrated the rates of in-patient care and critical care to be lower than the ones predicted by Ferguson et al. for the UK (See Supplement Information, Table S1.2). Our results show that the demand on ICU beds can be reduced not only by a suppression strategy as successfully used in China, but also by a mitigation strategy. However, deaths cannot be effectively prevented in mitigation scenarios as many of those would occur independent of ICU demands. Of note, for the modes of death possible in our model (see Supplementary Information, Table S1.3), the ICU demand can be maintained at a lower level while deaths rise, which appears to align well to the Swedish reported data including a substantial number of deaths occurring outside hospitals.

Our analyses address the impacts from COVID-19 on the health-care demand, deaths, and direct healthcare costs in Sweden in relation to different public health interventions. As such it is in line with the assessment of the European Center for Disease Prevention and Control regarding the COVID-19 pandemic. We find that the direct health-care related costs are substantial ranging between 4 and 20 billion SEK dependent on the scenario. We thus showed that more stringent mitigation or suppression efforts yield larger direct health care cost reductions compard to less stringent mitigation or suppression, or the counterfactual scenario, with the maximum cost difference estimating 17 billion SEK. These cost estimates are likely underestimating the true costs because the estimates are only measuring the direct costs per patient per day during normal health-care demand in 2018. Further on, the costs of the health sector would need to be balanced against the cost of the economy as a whole. The estimates here do not capture impacts within the healthcare from other acute health-problems for which treatment is down prioritized or postponed due to the acute situation of the epidemic. It does further not consider economic impacts beyond the health sector.

We also did not consider the role of seasonality. Underreporting, or asymptomatic transmission, can be a driver of herd-immunity. Preliminary findings from Iceland found no more than 50% asymptomatic carriers.(21) Ferguson et al. estimated an CFR around 1.6% and and IFR of around 0.8% based on an 50% asymptomatic rate. Our study found higher rates of mild and asymptomatic transmission fitted better with the Swedish data and estimates slightly lower IFR’s. Preliminary reports from New York and Germany indicates this may be the case, but further investigation with more robust estimation of antibody prevalence and infection fatality rates are needed.

Our study supports that public health interventions such as social distancing combined with shielding of older persons, even without a strict lock-down, can protect the health care system by not exceeding the ICU capacities, but resulted in more cases including deaths compared to neighbouring countries with similar population densities that introduced more stringent lock-down measures.

## Data Availability

The data can be accessed in correspondence with the authors.

## Data Availability

The data can be accessed in correspondence with the authors.

## Supplementary Information

### Supplementary Information 1: Compartmental model and parameterization

To account for time delays in the spatial spread of SARS-CoV-2 over the large geographical ranges in Sweden, we set up a spatial compartmental model. We distinguish between local (municipality) and global (Sweden) processes. The effects of local contact structures are assumed to be well described by the law of mass action; at local scales we assume a well-mixed contact-structure. The effects of global contact structures are assumed to be well described by a radiation model,(1) which gives rise to time delays in the spatial progression of infections over Sweden.

#### Extended SEIR-model

We apply an age-structured SEIR-based compartmental model for each municipality. In each municipality *i,* we account for all individuals that are susceptible *S_i_;* latent (exposed) *E_i_;* infectious but not going into healthcare *I_i_*; infectious and going into some form of hospital care *J_i_*; in healthcare *H_i_*; in intensive care *C_i_;* recovering in healthcare after critical care 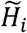; dead due to SARS-CoV-2 infection *D_i_*; still infected but not transmitting to others e.g., isolated or otherwise removed so to not potentially transmit 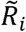, or, recovered *R_i_.* Each respective variable is age-structured (i.e., vectors with age-specific component values). We account for three age-classes, *a =* (0 − 59,60 − 79,80 +) years. The compartmental model, with dot-notation for time derivatives, can then be written

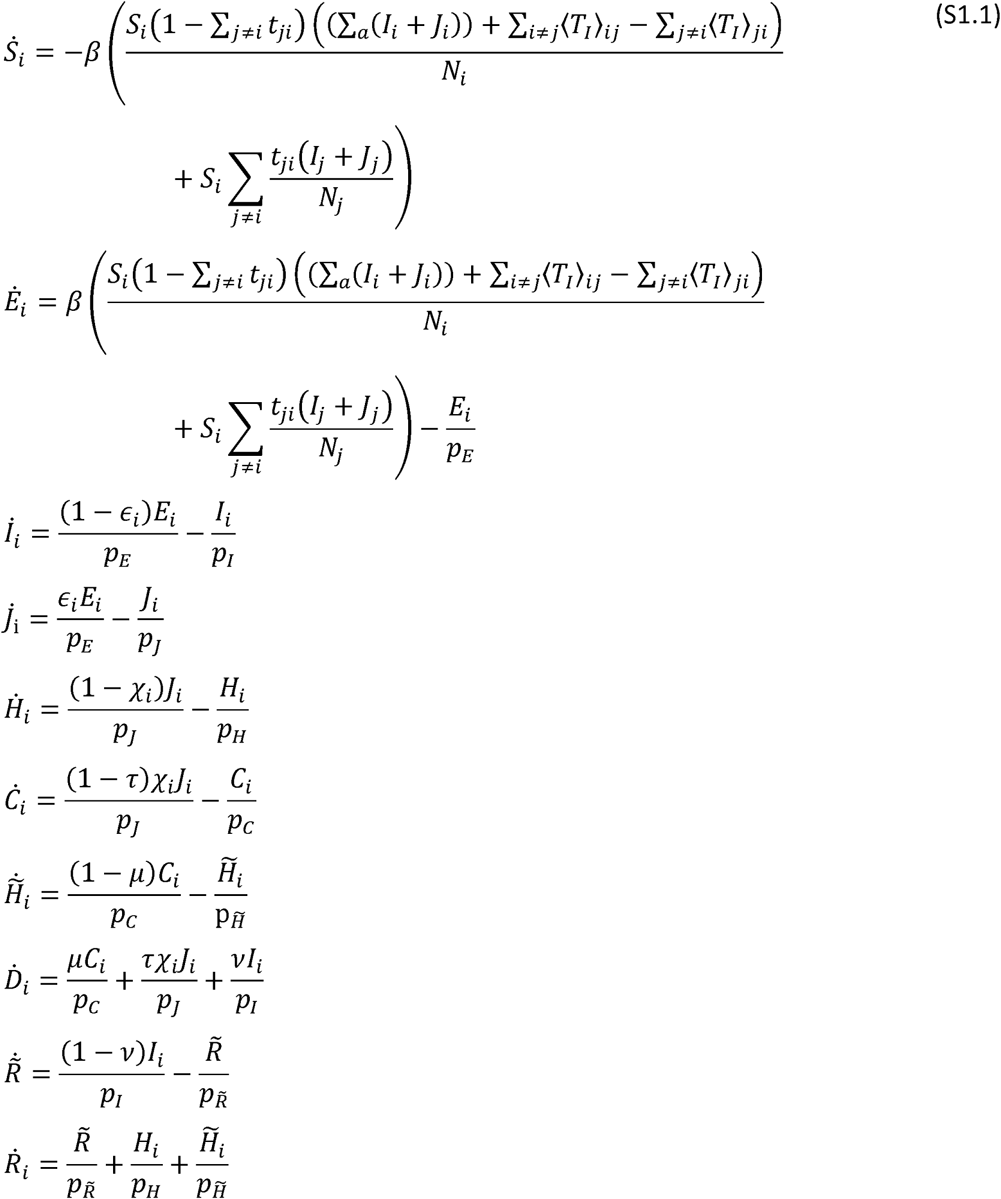

where 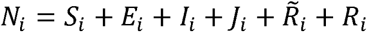 and see table S1.1 for model parameterization. Infection are carried between municipalities, and 〈*T_I_*〉_*ij*_ denotes the number per day of infected individuals that are resident to the *j*th municipality and are visiting the *i*th municipality; *S_i_*∑_*j≠i*_ *t_ji_* denotes the number per day of susceptible individuals that are resident to the *i*th municipality and are visiting other municipalities. This should be seen as daily averages.

#### Radiation model

These mobility rates are given by a radiation model^1^, where we used the time dependent rate scaling *α(t),* with 0.01 as the baseline, i.e., the counter-scenario of inter-municipality travel-reductions. The radiation model for the average number (denoted by angle-brackets) of travellers per day of *X_i_*, from municipality *i* to municipality *j,* can be written 〈*T_X_*〉*_ji_* = *X_i_t_ji_*, with the per person travel-probability

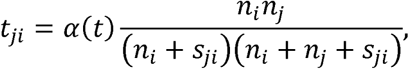

where *n_i_* is the number of citizens in the municipality *i*; where *S_ji_* is the total population size within a circle with a radius equal to the distance between two municipalities *i* and *j.* Note that *t* without subscript denotes time. See table S1.1 for *α*(*t*).

#### Geographic and demographic data

Population data were collected from the Statistics Sweden and the demographical geographical statistical units’ database. The database provides population data in 5-year age categories for almost 6000 administrative areas in Sweden and was compiled by the end of 2018. We aggregated this population data to the municipal level and into three age-groups: 0–59, 60–79 and older than 79 (i.e., 80+) years.

The geographical centroid coordinates (latitude, longitude) of municipalities were derived from shape-file data by using the R software libraries sp, rgdal, rgeos and foreign. These coordinates were used to calcluate the distances between each and all of the 290 Swedish municipalities. A distance matrix was constructed, and used to derive inter-municipality travel rates given by the radiation model.

#### Heterogeneous basic reproduction number (R_0_)

For the SEIR-model formality without vital dynamics, *R*_0_ is equal to *β* times the infectious period. As our model extends in some aspects from the stylized SEIR-model, *R*_0_ in our model is approximately given by this product. *R*_0_ depends on the contact-rate and infectious period. As both of these parameters are age-dependent, and that each municipality has unique age-distributions, our model accounts for a heterogeneous *R*_0_ which vary between municipalities. The age-dependence for contact rates comes from our assumptions on th within-municipality contact-structure (Supplementary Information 2), and relate directly to our modelled and presented scenarios. The age-dependence for infectious period (i.e., in this context, the number of days an infected individual can infect other individuals), arises here indirectly from that the length of the infectious period differs between those individuals that go into healthcare (and being practically isolated) and those that do not go into heathcare, and the proportion among infected individuals in ether of these two groups is age-dependent. Accordingly, *R*_0_ per municipality, without counter measures (i.e., for scenario a), varies within the interval 2.73 and 4.55. The low extreme would imply a three-days infectious period if all infected individuals go into healthcare, and the upper extreme would imply a five-days infectious period if no infected individuals go into health care. As the proportion of infected individuals going into healthcare is never zero nor one (Table S1.2), we know that none of the extremes in the interval becomes realized, yet that *R*_0_ is within the interval [2.73,4.55].

#### Numerical analysis and calibration procedure

The equation system (eq. S1.1) was solved in Matlab for respective variables by implementing the ordinary differential-equation solver ode45, with the initial condition that all individuals were susceptible except for 1/30000 of the population in Region Stockholm and 1/120000 of the population in all other Swedish Health-care regions (and municipalities), respectively.

The solved variables could accordingly be compared against empirical data on deaths, ICU load, healthcare load and virus-prevalence data. The model parameterization was iteratively calibrated agaist these data with the objective to narrow down parameter-values to fit the dynamics of one of the five scenarios to these data (for R^2^- and mse-values, see Table 1 in the main text). Data on COVID-19 deaths- and hospitalization were obtained through various online data aggregating services, (3,4) Data regarding healthcare load, ICU occupancy, and confirmed deaths were reported directly from the healthcare regions as well as the Swedish Public-Health Agency. Data collection was restricted up to and including the 17^th^ of april to account for lag in reporting. The virus carriage data were fetched from a study performed by the Swedish Public-Health Agency.(5)

#### Deriving Swedish COVID-19 hospitalization and death frequencies

Drawing on age-structured data^2^ on the proportion of reported cases requiring inpatient care (Table S1.2), we derived the corresponding *proportion of infected cases requiring hospitalization* (*∈_i_*) for any municipality *i* and for age-classes *a =* (0 − 59,60 − 79,80+) by taking a weighted average for each age-class in *a* and multiplying by 1/6 (obtained by calibration to Swedish data) We also derived the *proportion of hospitalized cases requiring intensive care* by the weighted averaging. Note that we further assumed a 85% and a 10% intensive-care triage for the ages 60–79 and 80+ years, respectively.

Mortality risk among the group of individuals receiving intensive care (i.e., *μ_i_*; Table S1.1) was derived by forming weighted averages of mortality risk for age-groups 0–59, 60–79 and 80+, based on the observed mortality risk in Italy.(6) We assume that the mortality risk in inpatient care outside the ICU is zero (a critically ill patient would be transferred to the ICU), and that the mortality risk among the group of individuals that is not admitted to the healthcare system (for different reasons), is proportional by a factor 0.008 (estimated by calibration to Swedish data) to the ICU mortality-risk distribution. We assume that patients that are not prioritized for care in the ICU based on the medical ethical principles set up in Sweden the pandemic will soon die from COVID-19.(7) The resulting country-level mortality-risk in Sweden for infected individuals in age-groups 0–59, 60–79 and 80+ is presented in Table S1.3.

#### Scenarios and within-municipality contact structures

Table S1.4 provides overall contact-rate scaling *ĉ* within and between age groups. They can be seen as reduction coefficients to *R*_0_ as a result of physical distancing. They can be interpreted as probabilities of exposure to potential contacts (e.g., the probability of going to public places, or equivalent, and not keeping safe distancing) times the contact-rate *q* given exposure to potential contacts (i.e., the contact-rate at public places), where the latter was normalized so that the overall contact-rate scaling equals to 1 without any suppression measures. Specifically, *ĉ* is equal to *p_a_p_b_q* for any age groups *a* and *b,* where *p_a_* and *p_b_* are probabilities that any individual from age group *a* and *b,* respectively, expose themselves to situations where potential contacts are possible. This can be generally expressed by a symmetrical “contact-matrix”

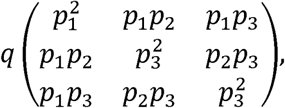

which is provided in Table S1.4 paremerizised for each respective scenarios a to e (e.g., figure 3 in main text), where any entries left of the main diagonal was omitted due to symmetry. Table S1.4 presents exactly the modelled scenarios a to e, with the only exception that scenario d and e in addition account for increased isolation of infectious individuals; modelled by reductions, 34% and 40%, respectively, in the infectious period among individuals not going into healthcare. The different scenarios are generally described by contact rates according to:

a. no changes in contact rates (baseline model);
b. 25% reduction in contacts in ages 0–59 years and 50% reduction contact in ages 60+ years.
c. 25% reduction in contacts in ages 0–59 years and 75% reduction in ages 60+ years;
d. 50% reduction in contacts in ages 0–59 years and 90% reduction in ages 60+ years and reduction of infectious period to 3.3 days among the general infected population by home isolation;
e. 50% reduction in contacts in ages 0–59 years and 90% reduction in ages 60+ years and further reduction of infectious period to 3.0 days among the general infected population by home isolation.

See Table S1.4 for the between age group contact rate reductions for scenario (a)-(e).

**Table S1.1.**
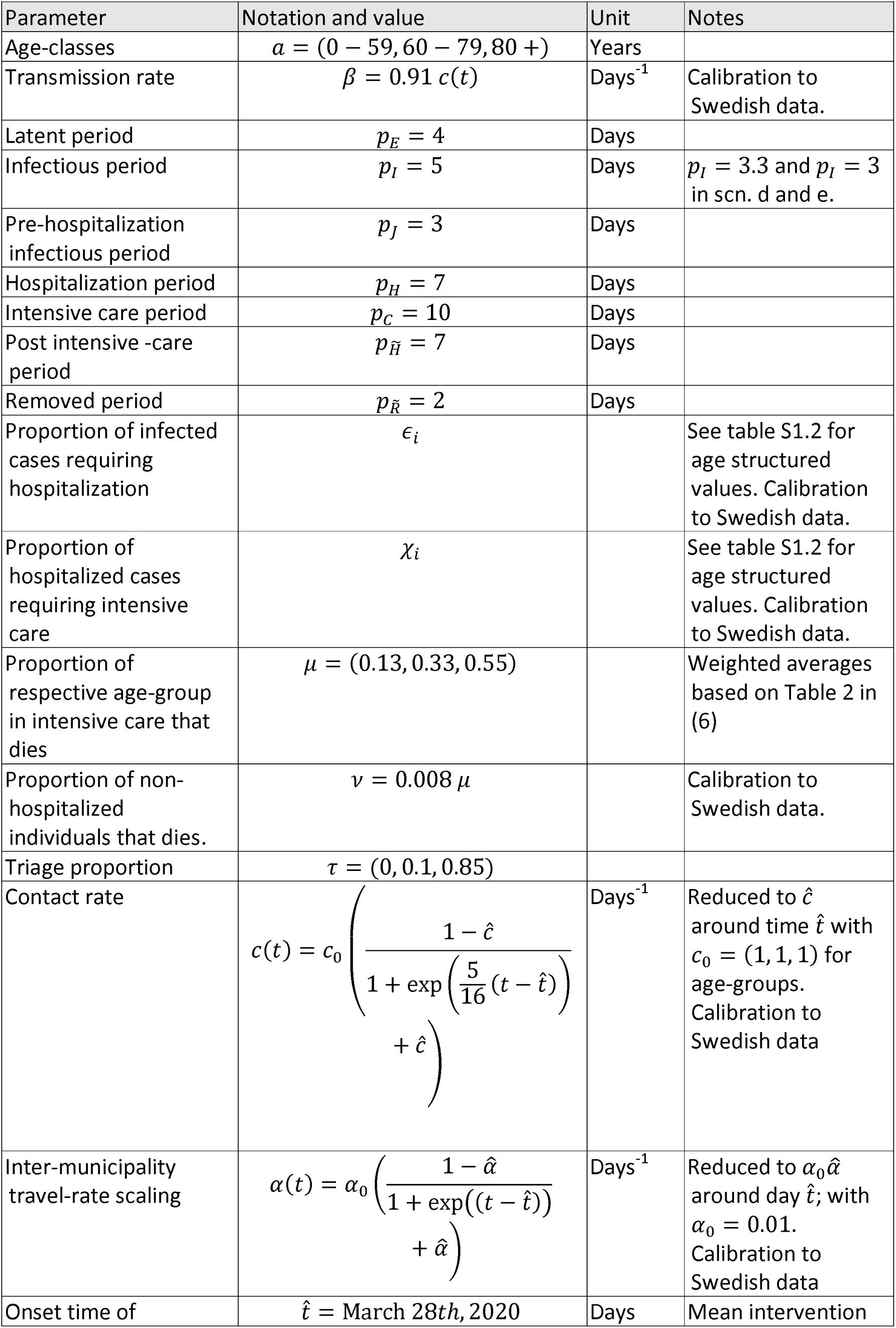
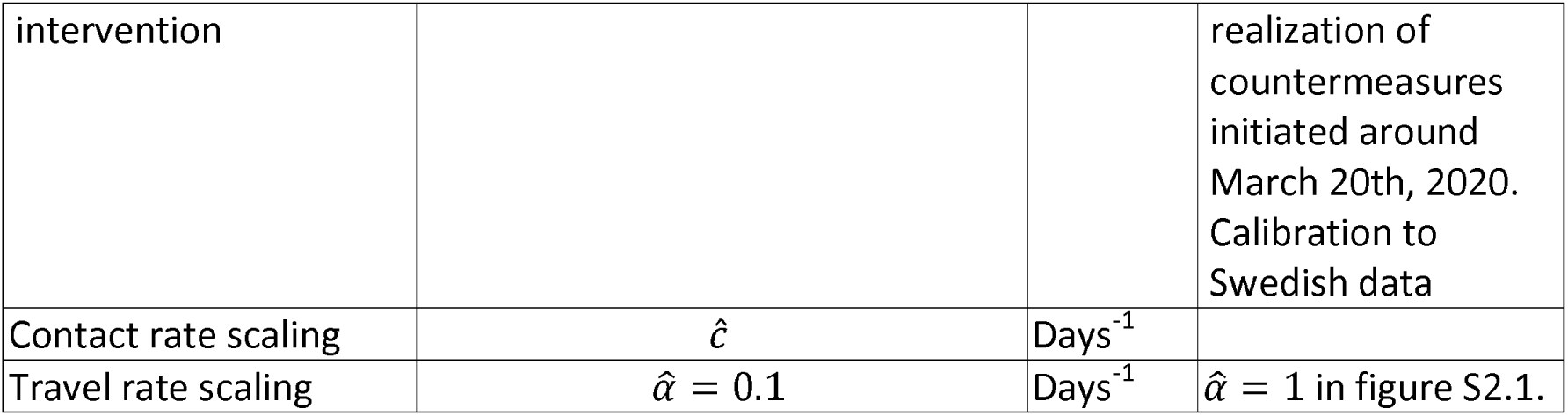
Parameters in equation system S1.1 and their respective values.

**Table S1.2.** Conditional risks for in-patient care and intensive care

| **Table S1.2.** Conditional risks for in-patient care and intensive care |  |  |  |  |
| --- | --- | --- | --- | --- |
| Age-class<br>(years) | From Ferguson<sup>2</sup> |  | Calibrated to Sweden & Stockholm |  |
| Age-class<br>(years) | Percent of<br>reported cases<br>requiring<br>inpatient care | Percent of in-<br>patient cases<br>requiring intensive<br>care | Percent of<br>infected<br>persons going<br>into inpatient<br>care | Percent of in-<br>patient cases<br>requiring<br>intensive care |
| 0-9 | 0.1 | 5.0 | 0.02 | 5.0 |
| 10-19 | 0.3 | 5.0 | 0.05 | 5.0 |
| 20-29 | 1.2 | 5.0 | 0.20 | 5.0 |
| 30-39 | 3.2 | 5.0 | 0.53 | 5.0 |
| 40-49 | 4.9 | 6.3 | 0.82 | 6.3 |
| 50-59 | 10.2 | 12.2 | 1.70 | 12.2 |
| 60-69 | 16.6 | 27.4 | 2.77 | 27.4 |
| 70-79 | 24.3 | 43.2 | 4.05 | 43.2 |
| 80+ | 27.3 | 70.9 | 4.55 | 70.9 |

**Table S1.3.** Mortality risk among infected populations (%)

| **Table S1.3.** Mortality risk among infected populations (%) |  |  |  |
| --- | --- | --- | --- |
| Age-class<br>(years) | In intensive care | After triage prior to<br>intensive care | Outside of hospital-<br>care |
| 0-59 | 0.0052 | 0 | 0.11 |
| 60-79 | 0.35 | 0.12 | 0.26 |
| 80+ | 0.26 | 2.74 | 0.42 |

**Table S1.4:**
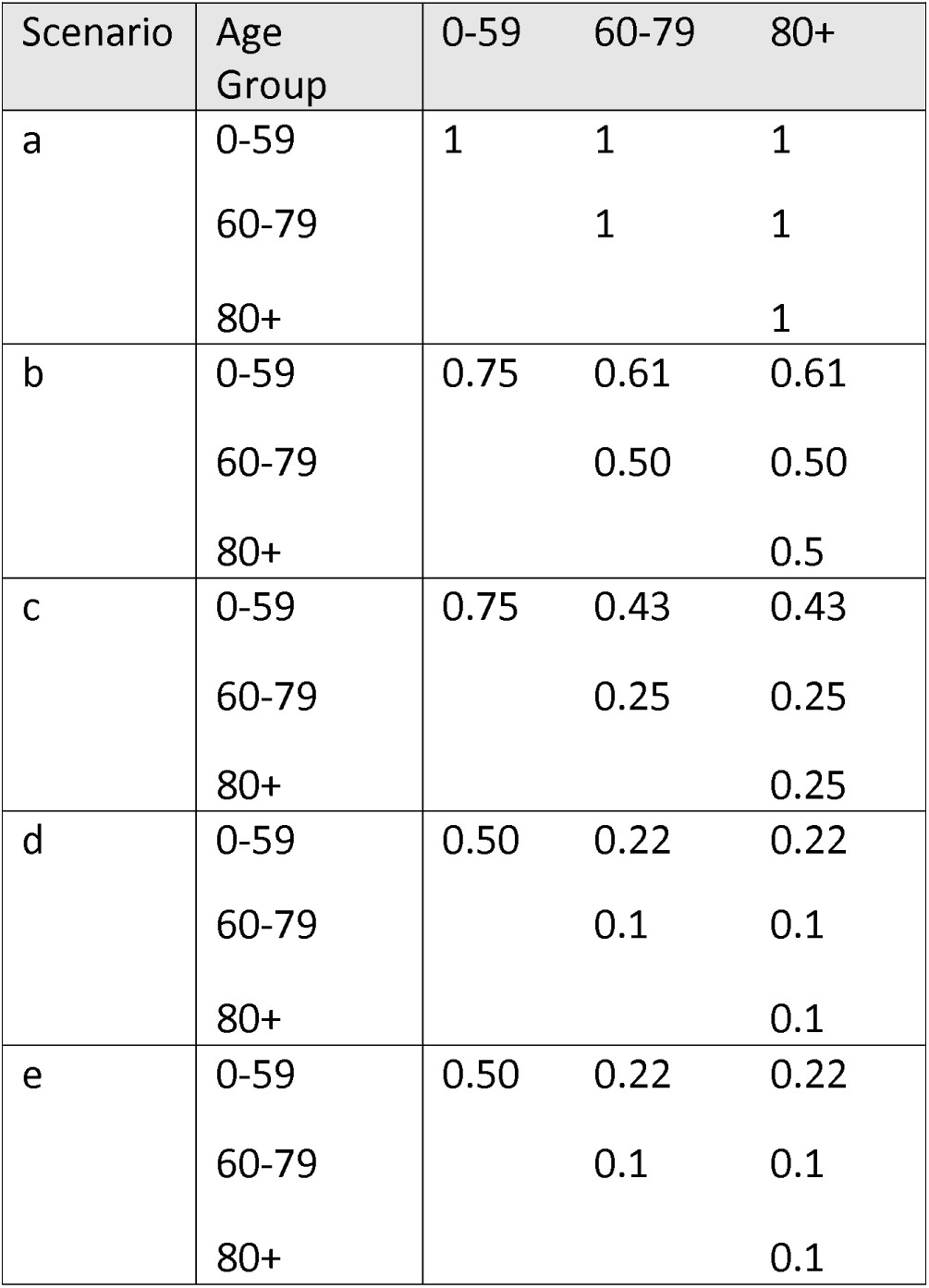
Contact rate scaling, i.e., *ĉ* (Table S1.1), by scenarios (a) to (e).

| **Table S1.4:** Contact rate scaling, i.e., $\hat{c}$ (Table S1.1), by scenarios (a) to (e). | | | | |
| --- | --- | --- | --- | --- |
| Scenario | Age Group | 0-59 | 60-79 | 80+ |
| a | 0-59 | 1 | 1 | 1 |
|  | 60-79 |  | 1 | 1 |
|  | 80+ |  |  | 1 |
| b | 0-59 | 0.75 | 0.61 | 0.61 |
|  | 60-79 |  | 0.50 | 0.50 |
|  | 80+ |  |  | 0.5 |
| c | 0-59 | 0.75 | 0.43 | 0.43 |
|  | 60-79 |  | 0.25 | 0.25 |
|  | 80+ |  |  | 0.25 |
| d | 0-59 | 0.50 | 0.22 | 0.22 |
|  | 60-79 |  | 0.1 | 0.1 |
|  | 80+ |  |  | 0.1 |
| e | 0-59 | 0.50 | 0.22 | 0.22 |
|  | 60-79 |  | 0.1 | 0.1 |
|  | 80+ |  |  | 0.1 |

### Supplementary Information 2: Scenario with no reduction in inter-municipality travel rates

Figure S2.1 shows the results on ICU load should there be no reduction of inter-municipality travel rates, and with other things equal to the model parameterization for which Figure 3 in the main text is the result.

**Figure S2.1.**
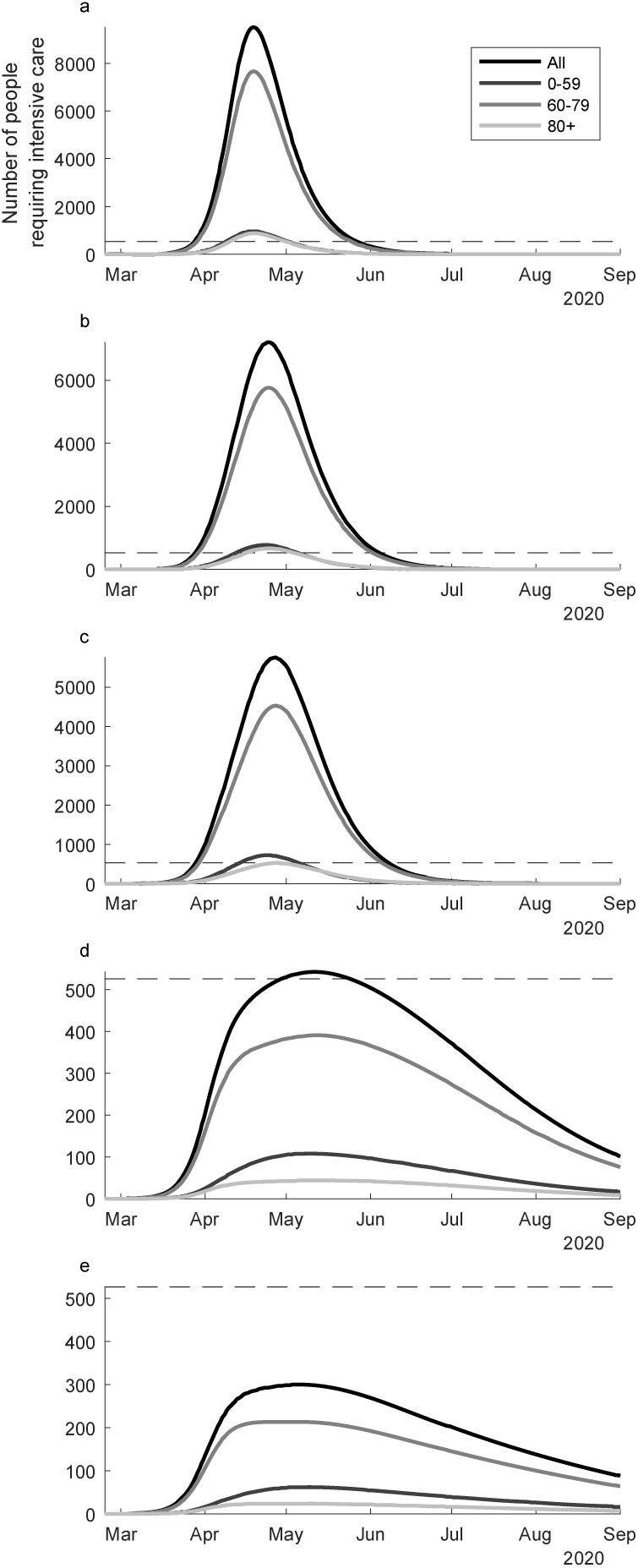
Scenarios of ICU demand simulated based upon higher mobility between municipalities as described by the radiation model.

### Supplementary Information 3: Analysis of excess mortality in Stockholm

Using mortality data from Statistics Sweden (8) and officially confirmed COVID-19 mortality data reported by the 21 regions of Sweden collected up until the 11^th^ of April, 2020,(3) the excess mortality in response to reported COVID-19 deaths was estimated for the region of Stockholm using a linear regression model (see Figure S3.1). The outcome of this model was the daily total number of deaths in all-causes, subtracting the number of deaths observed in COVID-19. The number of deaths was assumed well described by a normal distribution due to the higher frequency of events. The analysis adjusted for time trends using a factor variable for year and season and a factor variable for month. The daily COVID-19 deaths were smoothed using a moving average with a window of 7 days. The excess mortality was modelled as a linear function and estimated of a linear increase of 0.40 (95% CI = 0.24, 0.57) increase in all-cause non-COVID-19 mortality for every COVID-19 death.

**Figure S3.1.**
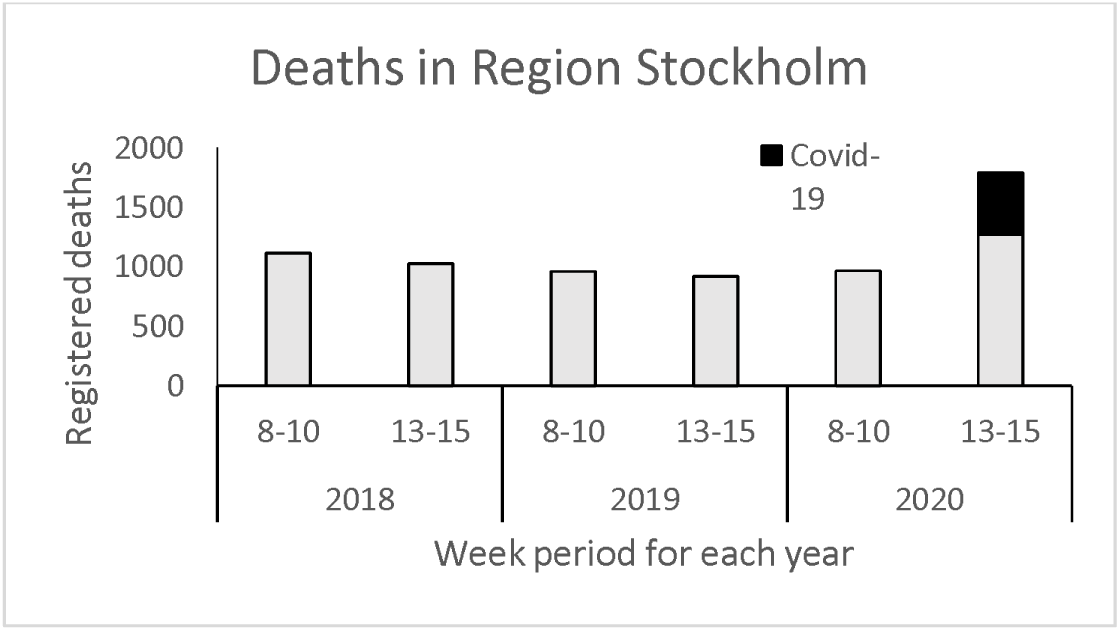
Deaths for weeks 8–10 and 13–15 in year 2018, 2019 and 2020 for the region of Stockholm, Sweden.

### Supplementary Information 4: Cost estimates

The cost estimates were retrieved using Cost per Patient (CPP) database that reports costs for every individual care event. The database use Diagnose Related Group (DRG) to group the care events to be able to report costs that describe well the average costs at a group level. The database provides average time in care and the average costs per care event.(9)

#### Average treatment costs calculation for inpatient care

To account the costs per day in care, we used costs reported under following diagnose codes S.40 Virus infection; Main diagnoses: B.349 Virus infection, unspecified; J.108 Influenza due to other identified influenza virus with other manifestation. Average cost per day was calculated dividing the total cost with average number of days spent in care.

#### Average treatment costs calculation for ICU care

To calculate the ICU care all costs reported under respiratory diseases (D.20) receiving invasive ventilation treatment were extracted from the database and divided by average number of days for treatment to calculate the average cost per day.

For both inpatient and ICU care cost estimates, the total average cost for all age groups reported in 2018 was used. All cost estimates were adjusted for consumer price index and reported in 2020 SEK value. Cumulative patient days were multiplied with the average cost estimates for inpatient care and ICU care.

The Swedish Intensive Care Register Yearly Report 2018 estimates that an average cost per day in intensive care is from 50 000 – 80 000 SEK, which is a higher estimate that the one we are using here.(10) The lower estimate was chosen to avoid overestimations of direct costs, Notably, the COVID-19 costs for a proportion patients receiving extracorporeal membrane oxygenation treatment was not included here.

